# Individual-level response adaptive crossover trial design for epilepsy: structure and simulation

**DOI:** 10.1101/2020.10.09.20210286

**Authors:** Wesley T. Kerr, Xingruo Zhang, John M. Stern

**Affiliations:** Department of Neurology, David Geffen School of Medicine at the University of California, Los Angeles, Los Angeles, California, USA; Department of Internal Medicine, Eisenhower Health, Rancho Mirage, California, USA

**Keywords:** epilepsy, clinical trial simulation, medication response, seizure frequency, Poisson process

## Abstract

Trials of antiseizure medications involve static group assignments for treatments with pre-specified durations. We propose a response-adaptive crossover design using basic statistical assumptions regarding both seizure count and duration of treatment to determine when a participant can change group assignment. We modelled seizure frequency as a Poisson process and estimated the likelihood that seizure frequency had decreased by 50% compares to baseline using both a Bayesian and maximum likelihood approach. We simulated trials to estimate the influence of this design on statistical power and observation duration with each treatment. For patients with 9 baseline seizures in 4 weeks who had no change in seizure frequency, the simulation identified non-response in a median of 16 days. The response-adaptive crossover design resulted in a modest increase in statistical power to identify an effective treatment while maximizing the time in a group producing a response. Only 8% of participants remained in the placebo group for all 90 days of the simulated trials. These example theoretical results can provide quantitative guidance regarding objective criteria to determine non-response in real-time during a controlled clinical trial without revealing the assigned treatment. Implementing a response-adaptive crossover design may both improve statistical power while minimizing participant risk.

## 1. Introduction

Clinical trials for treatments of epilepsy examine treatments that may reduce the frequency of epileptic seizures, ideally to zero. Effect sizes in these trials are limited by waiting to observe potentially infrequent seizures and uncertainty in the real-time criterion for response to treatment. Response to treatment is based on the flawed measure of participant-reported seizure frequency, as there is no objective biomarker for response ^1^.

Worsening or non-response to treatment results in risks including the immediate physical injury during and after the seizure, loss of independence from driving, employment and other restrictions, and the social stigma of seizures. In fact, participants who were assigned to the placebo arm of clinical trials have been shown to have a 6.1 times increased risk of sudden unexpected death in epilepsy (SUDEP) ^2^. In addition, there are direct financial costs both to the participant and trial administration to continue participants whose assigned treatment is not producing a positive response. Therefore, there have been substantial discussions regarding how exposure to placebo or ineffective treatments can be minimized ^3^. Implementation of response-adaptive randomization may address many of those concerns ^4; 5^.

To illustrate this approach and advance the optimization of response-adaptive design, we propose a statistical criterion to define how long a participant receives a treatment based both upon the number of seizures that occurred and the length of time during which those seizures occurred. This builds upon the previously proposed time to the pre-randomization seizure count (PSC), which expects that this time would be lengthened by effective treatment ^6^. The exact statistical process that models seizure occurrences has not been determined (see discussion and ^7; 8^). For illustration, we simplify this emerging literature by modeling seizures as a Poisson process and defining non-response as less than a 50% reduction in seizure frequency, with a false positive rate (α) of 5%. After individual-level response is determined, the participant is switched to another treatment arm. If the participant has already been in all treatment arms, participation is either terminated or the re-assignment is to the treatment arm where they had the lowest seizure frequency for the remainder of the trial.

## 2. Materials and Methods

The participant’s journey through this individual-level response adaptive cross-over randomization involves two steps (Figure 1). The first step is the real-time criterion to determine when a participant would be unlikely to respond to the assigned treatment. The second step is the process of response-adaptive crossover trial design to select subsequent treatments after response has been determined.

**Figure 1.**
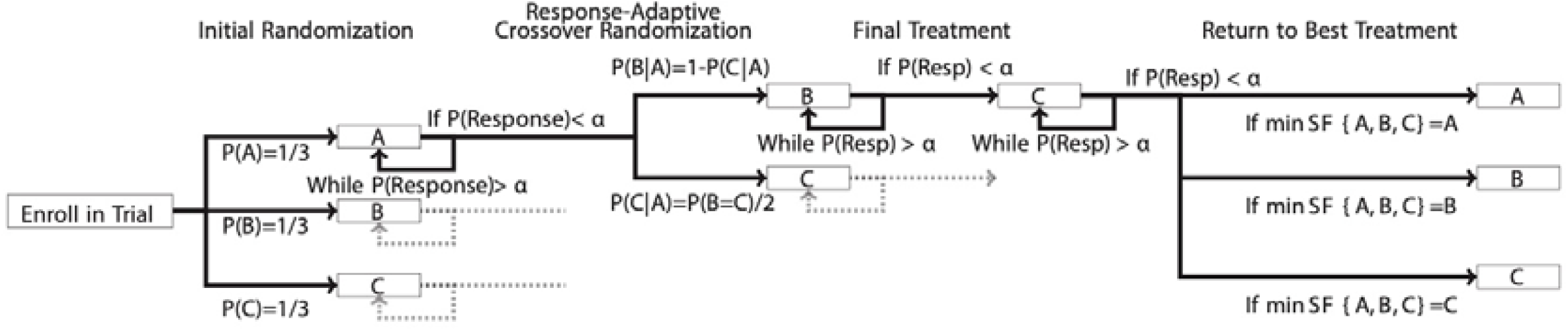
Sparse Flow sheet of the proposed Response Adaptive Crossover Trial for three independent treatments: A, B and C. The dashed line indicates that each type of branch occurs for all other arms but is omitted for clarity. Step 1) Initial balanced randomization. Step 2) If the probability of response to initial treatment, P(Response) or P(Resp), is less than α (e.g. 5%), then response-adaptive randomization based on statistical comparison of other treatments. Step 3) If probability of response to second treatment is less than α, then assign to last treatment. Step 4) If probability of response to third treatment is less than α, then assign to the treatment with the lowest observed seizure frequency (SF) for that individual.

### 2.1 Determination of Individual-Level Response

This manuscript utilizes statistical theory to determine the minimum number of seizures reported by an individual participant at any point within a clinical trial that would make it unlikely that the participant’s SF has halved during the treatment period. While the structure of a clinical trial can vary, we specifically address trials with an initial 28-day baseline, or pre-randomization, monitoring period, followed by randomization into the assigned treatment for 90-days. The proportionality of Poisson processes allows our method generalizes to any length of baseline or treatment period.

The following will demonstrate the statistical basis by which we determine the probability that a patient has either failed or responded to an anti-seizure medication (ASM) based on real-time reporting of their seizures during a clinical trial. This is organized in the following sections:

(2.1.1) Stating the problem based on statistical assumptions and distributions
(2.1.2) Illustrating the framework of the solution using conventional hypothesis testing
(2.1.3) Incorporating uncertainty of estimation into the solution using a Bayesian approach

We first address the question of how to determine the likelihood that the patient has failed a medication. Subsequently, we address the probability of falsely identifying a patient as a non-responder if the threshold of experiencing more than half the baseline seizures is used.

#### 2.1.1 Statistical Problem Statement and Assumptions

First, we model the statistical process that generates seizure counts, *N*, over a certain period of time, *t*, as Poisson processes in which the following assumptions are made:

- The probability of a seizure during a time interval is proportional to the length of that time interval.
- Two seizures cannot occur simultaneously.
- The probability that a seizure occurs does not change over time within the clinical trial unless a treatment change is made.
- The probability that a seizure occurs during one-time interval is independent of if a seizure occurs during any non-overlapping time interval.

While most of these principles are theoretically true for seizures, real-world studies have shown deviation from this theoretical ideal ^9-11^. We chose to use a Poisson process model because both the number of seizures and time in which these seizures occurred can be used to develop more efficient criteria for response. As we discuss below, this approximation of probability of individual-level response is only as good as the Poisson approximations hold true for each individual’s seizure pattern. In other words, this Poisson assumption may not be accurate, but it may be useful.

Others have suggested that a negative binomial distribution resulted as much as 15% reduction in residual error when modeling seizure counts ^7^. While the negative binomial may model seizure count more closely, the difference between the Poisson and negative binomial does not influence the utility of our proposed trial design. We chose to use a Poisson to be able to make clear statements regarding the assumptions of the model and determine the shape of the Bayesian priors in 2.1.2 and derive the analytical formulae in 2.1.3.

Based on the properties of a Poisson process, we then determine that the probability that a patient has *n* seizures during time interval *t* is given by:

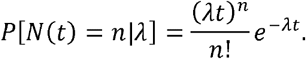

In a clinical trial with a baseline period, we can find the maximum likelihood estimate of the baseline *λ*, denoted as *λ*_0_, for each patient with the following formula:

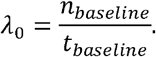

In a clinical trial where we are assessing if a patient responds to an antiseizure mediation (ASM), we define a response as a 50% reduction in seizure frequency. In our notation, that would be equivalent to halving *λ*_0_. Therefore, in the treatment phase, we evaluate, given our reported seizure count, *n*_*t*_, the probability that *λ*_*t*_ is less than or equal to half of *λ*_0_.

The threshold of response of halving of the SF is based on European Union convention for the reporting of trials in seizures ^12^. These thresholds can be modified based on the individual trial design. Reasonable choices could include 25% reduction, non-inferiority of the baseline, or less than doubling of the seizure frequency. The latter threshold, similarly, has been used as a criterion for exit in trials before ^13-15^.

#### 2.1.2 Framework of the Solution

Therefore, for a given seizure count during the treatment period, *n*_*t*_, we determine the probability that seizure count, or higher, would occur if the rate of seizures had at least halved. This corresponds to a null hypothesis that the seizure rate is *λ*_*t*_ ≤ *λ*_0_/2 and a one-tailed alternative hypothesis that *λ*_*t*_ > *λ*_0_/2. We write the probabilistic expression as:

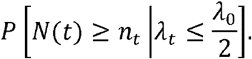

If we take the worst case scenario that the seizure rate had just halved, we recognize this as the right sided cumulative distribution function of the Poisson distribution, which is the upper incomplete gamma function, 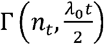. This logic is illustrated in the following expression:

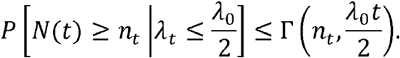

Therefore, based on the reported seizure frequency during the baseline period, we can create a table of situations where this probability is less than 5%, suggesting that the null hypothesis that seizure frequency has been halved is violated with false positive rate, α, of 5% (Table 1).

**Table 1.** The minimum number of seizures to be observed in a given number of days for the probability of 50% response to be less than 5% using the Bayesian approach. Abbreviations: seizure (Sz), day (d), week (w).

| Baseline 28d | | Minimum number of sz in X days to have not responded with $\alpha<0.05$ | | | | | | | | | | | | | |
| --- | --- | --- | --- | --- | --- | --- | --- | --- | --- | --- | --- | --- | --- | --- | --- |
| Sz Count | Sz Rate | 1 | 2 | 3 | 4 | 5 | 6 | 7 | 14 | 21 | 28 | 30 | 60 | 90 |  |
| 100 | 3.6/d | 4 | 7 | 9 | 12 | 14 | 16 | 19 | 34 | 50 | 64 | 69 | 132 | 194 |  |
| 56 | 2/d | 3 | 5 | 6 | 8 | 9 | 11 | 12 | 21 | 31 | 40 | 42 | 80 | 117 |  |
| 28 | 1/d | 2 | 3 | 4 | 5 | 6 | 6 | 7 | 12 | 17 | 22 | 24 | 44 | 65 |  |
| 14 | 1/2d | 1 | 2 | 2 | 3 | 3 | 4 | 4 | 7 | 10 | 13 | 13 | 26 | 38 |  |
| 9 | 1/3.1d | 1 | 1 | 2 | 2 | 3 | 3 | 3 | 6 | 8 | 10 | 11 | 20 | 28 |  |
| 4 | 1/w | 1 | 1 | 1 | 1 | 2 | 2 | 2 | 3 | 5 | 6 | 6 | 12 | 17 |  |
| 2 | 1/2w | 1 | 1 | 1 | 1 | 1 | 1 | 1 | 2 | 3 | 4 | 5 | 9 | 12 |  |
| 1 | 1/4w | 1 | 1 | 1 | 1 | 1 | 1 | 1 | 2 | 2 | 3 | 4 | 6 | 9 |  |

As a point of discussion, if the actual new seizure rate, *λ*_*t*_, was less than half of *λ*_0_/2 then the actual seizure rate would be lower, but the number of seizures that needed to be observed prior to determining that the patient had not responded to the new treatment would not change. In this situation, the probability of enough seizures occurring to reject the null hypothesis would be lower than 5% but, because we observe seizures instead of *λ*_*t*_, we would have to consider the worst-case scenario that *λ*_*t*_ = *λ*_0_/2.

Secondly, this same framework can be used to address how often patients with seizure frequencies less than half the baseline seizure frequency report seizure counts higher than half the baseline rate during the treatment period. This expression is as follows:

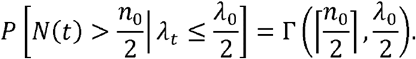

Where ⌈ and ⌉ are the ceiling operators and the left side of the equation models *N*(*t*) > *n*_0_/2 instead of *N*(*t*) ≥ *n*_*t*_.

These estimates focus on individual-level prediction instead of population-level prediction; therefore utilization of a population-level, mixed-effects model where some individuals are classified as responders and some are classified as nonresponders is not appropriate ^7; 10; 16; 17^. Mixed-effects models hypothesize that the population is a mix of non-responders with no change in seizure frequency plus responders with a common, group-level improvement in seizure frequency. Mixed-effects analysis reveals treatment assignments by comparing the distribution of placebo response to active-arm non-responders. Due to this focus on group-level response and unblinding, the individual-level likelihood that the participant is a responder is calculated after trial completion and is not determined in real-time during the trial.

#### 2.1.3 Incorporating statistical uncertainty

One complication behind the simple solution described above is that we utilized the maximum likelihood estimate of *λ*_0_ and *λ*_*t*_. Next, we consider the possibility that the actual estimated baseline seizure rate, 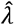, is different from this maximum likelihood estimate, and then repeat the process illustrated above. We incorporate this uncertainty by utilizing a weighted contribution of the possible options for *λ*_0_ where the weights are based upon the probability of a particular 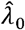 given the observed number of seizures in the baseline period, *n*_0_.

To estimate these probability-based weights, we use Bayes formula as below:

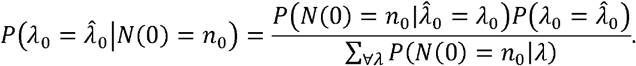

We estimate the prior distribution of 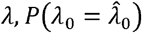, using a uniform distribution across all possible values of *λ*_0_ ranging from one seizure every 28 days to 24 seizures per day for 28 days in a row in steps of one seizure every 28 days. The shape of this probability-based weighting for example values of *n*_0_ is illustrated in Figure 1.

We also can use this structure to estimate the likelihood of each seizure rate, *λ*_*t*_, during the treatment phrase based on the reported seizures during the treatment phase, n_t_. In this way, we can assess the likelihood that 2*λ*_*t*_ ≤ *λ*_0_ using the following expression:

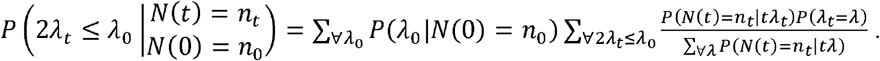

We include *t*in this expression because we are considering the number of seizures, *n*_*t*_, that occur in a certain number of days during the treatment period, which may be less than 28 days.

For the prior distribution of *λ*_*t*_, we again use the uniform distribution across the same range as prior. If we had assumed that *λ*_*t*_ was unchanged by treatment, this would bias our analysis towards concluding that the seizure rate during the treatment period, *λ*_*t*_, was unchanged from the baseline period, *λ*_0_.

We can determine the minimum *n*_*t*_for which this probability is less than 5% to determine the number of seizures needed to be observed in a specific treatment period to determine that the patient is unlikely to have responded to treatment. A table of these minimum seizure counts, *n*_*t*_, is displayed in Table 2.

**Table 2.** Characteristics of the duration of observation in 10,000 simulated participants with varying baseline seizure frequency (SF) and set change in SF with treatment. The Asterix indicates the minimum allowed duration of observation of 7 days. Abbreviations: Maximum Likelihood Estimate (MLE); Bayesian Posterior Probability (BPP); Upper Bound of the 95% empiric confidence interval (95% UB).

| Baseline SF |  | Change in SF: | MLE |  |  | BPP |  |  |
| --- | --- | --- | --- | --- | --- | --- | --- | --- |
|  |  |  | Half | No Change | Double | Half | No Change | Double |
| 4 | Days Observed (Median; 95% UB) |  | 90;90 | 27;90 | 7*;30 | 90;90 | 30;90 | 7*;37 |
|  | Completed 90 days (%) |  | 78 | 17 | 0.02 | 84 | 37 | 0.03 |
| 9 | Days Observed (Median; 95% UB) |  | 90;90 | 16;78 | 7*;16 | 90;90 | 16;90 | 7*;16 |
|  | Completed 90 days (%) |  | 80 | 4 | 0 | 87 | 13 | 0 |
| 28 | Days Observed (Median; 95% UB) |  | 90;90 | 8;27 | 7;7* | 90;90 | 8;31 | 7;7* |
|  | Completed 90 days (%) |  | 80 | 0 | 0 | 87 | 0.05 | 0 |

Secondly, to use this similar Bayesian framework for the question of falsely identifying pateints as non-responders when they report more than half as many seizures as they did during the baseline period. This corresponds to the following statistical expression:

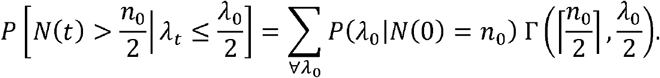

### 2.2 Simulated Trials

To illustrate the practical impact of this approach, we simulated a Poisson process for 10,000 participants with varying response to treatment from an 80% reduction in seizure frequency to a doubling of seizure frequency compared to the baseline seizure frequency. We chose 10,000 simulations based on the rule of thumb that 5% ordinal thresholds tend to be stable after 10,000 simulations. For clarity, our approach does not require that an individual trial have 10,000 participants. We illustrate these simulations for 4, 10, and 28 baseline seizures, chosen as low, median and high baseline rates based on previous recent trials ^13-15^. Using these simulations, we estimated the median and 95% confidence interval upper tail of the duration of treatment needed to determine non-response. To examine the false-positive rate for identifying non-responders, we varied the mandatory observation period from 1 to 45 days and simulated 100,000 participants who had a 50% reduction in seizure frequency.

### 2.3 Response-adaptive crossover randomization

After we have determined that the participant has not responded to their initial assigned treatment, we then determine which subsequent treatment to assign them to. In a two-arm trial, this would involve switching from active treatment to placebo or vice-versa.

In trials with multiple treatment arms, we can use other principles of response-adaptive randomization. Namely, due to the modeling above, we have an updating individual-level and therefore also population-level estimate of the likelihood each participant is responding to their assigned treatment. Based on current standards for clinical trials of seizures ^12^, we compared median percent reduction in seizure frequency of treatment relative to baseline. Due to potentially small sample size in some entries of the two-by-two contingency table of the median test, we used Fisher exact statistics as compared to Pearson’s chi-squared tests. While we could have weighted this comparison by the certainty of difference using the formulae in section 2.1 or the duration on the assigned treatment, we opted to maximally match the statistical techniques applied in canonical trials. The motivation for the median is that the theoretical ratio of λ_t_:λ_0_ - where λ is the parameter for a Poisson process - is not normally or t-distributed.

The subsequent treatment is probabilistically selected based on this population-level estimate of response. Specifically, the likelihood of selecting the potentially inferior treatment is proportional to the p-value of the population-level difference between the treatments. For example, in a trial with placebo and two doses of a single treatment, a participant who did not respond to the low dose active treatment could be randomized to placebo or high dose treatment, with a probability of 30% and 70%, respectively if the p-value of difference is 60%.

If the participant did not respond to this second treatment regimen, the participant would then be randomized to the last treatment protocol.

If a participant has not responded to any of the potential treatment protocols within the trial, there are two options: exit the trial or re-assignment to the treatment group that achieved the lowest seizure frequency for the duration of the trial’s observation period. This extended observation on the potentially most efficacious treatment allows for observation of long-term adverse effects on treatment.

As a trial progresses, there is increasing likelihood that the active treatment arms may be determined to be statistically superior to placebo with a false positive rate, α, of 5%. In this case, the above response-adaptive randomization scheme would have a less than 5% likelihood of assigning a participant to placebo if there were multiple treatment arms that the participant had not experienced. However, some participants would still be assigned placebo if the participant had not entered that arm. Instead of ending a trial early, an additional criterion could avoid this by eliminating the placebo option if pre-determined criteria are met. The remaining participants would receive one of the effective active treatments.

To illustrate this protocol through simulations, we consider 10,000 simulated trials with a placebo and two independent treatments, one of which causes a 25% reduction in seizure frequency and the other causes a 50% reduction in seizure frequency. To reduce the complexity of the simulation and have a simulation similar to the size of a typical RCT, we simulated 300 sequential participants per trial with baseline seizure frequency of 9 seizures per 28 days and use the BPP method for determination of individual-level non-response. We chose this seizure frequency as slightly higher than the median baseline seizure frequency in recent trials to benefit the utility of this design, but recruiting a participant population with this higher frequency is still reasonable ^13-15^. We did not use response-adaptive randomization to determine the initial treatment arm and we did not add an early termination criterion described in the paragraph above. We summarize these simulations by showing the empiric statistical power estimates and the length of time participants spent in each treatment arm using the BPP, the time to pre-randomization seizure count, and a canonical trial design.

While we simulate two independent treatments, we recognize that in most trials multiple treatment arms involve multiple doses of the same medication, leading to non-independent treatment arms. For example, if a participant was initially randomized to the high dose treatment and did not respond, it would be less likely that the participant would respond to the lower dose treatment. This level of complexity complicates the interpretation of our initial results; therefore, we deferred this consideration to future work.

## 3. Results

### 3.1 Estimating individual-level non-response

Table 1 and Supplementary Table 1 illustrate the minimum number of reported seizures during the treatment period that would be associated with a less than 5% chance that the participant’s SF had decreased by 50% using the BPP and MLE method of the baseline SF, respectively. The trial retention curves and statistics for 4, 10, and 28 baseline seizures in 28 days are illustrated in Figure 3 for BPP, Supplemental Figure 1 for MLE and both MLE and BPP are described in Supplemental Table 1. We found that requiring at least one week of observation substantially reduced false positive rates, and longer duration of mandatory observation reduced the rate of falsely identify participants as non-responders (Figure 4). The rate of falsely identified non-responders was independent of baseline seizure frequency. Refer to the online calculator linked below and supplemental text for more detailed information about other baseline seizure rates, durations of follow up and the minimum number of seizures to suggest no change or a doubling of seizure frequency.

**Figure 2.**
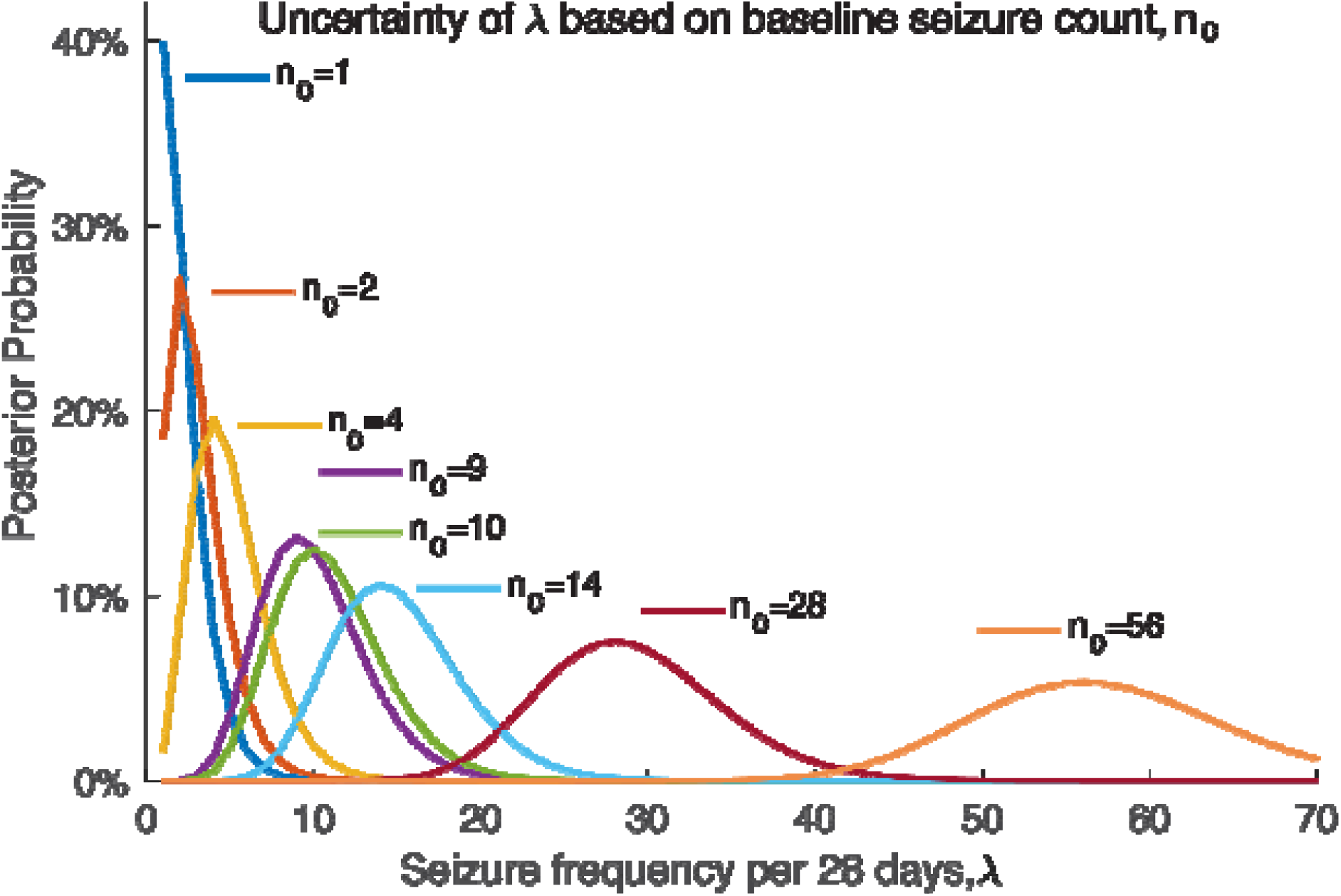
The spread of the Bayesian posterior probability of the baseline seizure frequency, λ_0_, based on the number of seizures reported in a 28-day period, n_0_.

**Figure 3.**
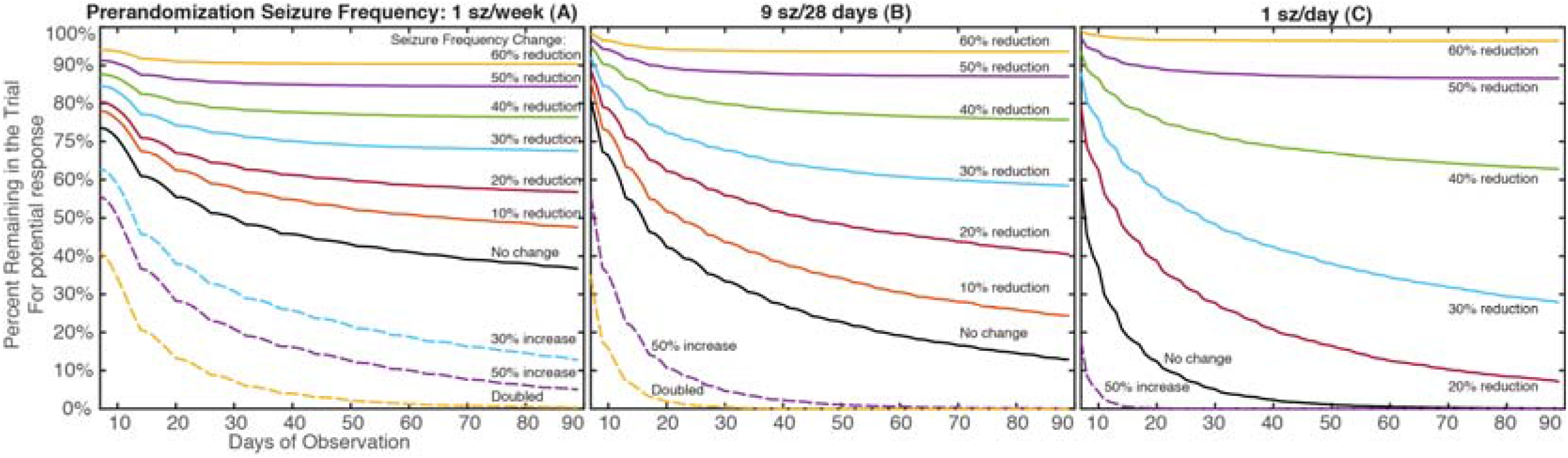
The pattern of retention within a 90-day trial when 10,000 simulated participants exited the trial when their interim seizure frequency (SF) suggested non-response for participants with 4 (A), 9 (B), and 28 (C) seizures in a 28-day pre-treatment period using the Bayesian approach. The minimum duration of retention was set to 7 days. The disjointed nature of the curves in 2A and less so in 2B reflect transition of the cutoff from one integer to the next highest integer.

**Figure 4.**
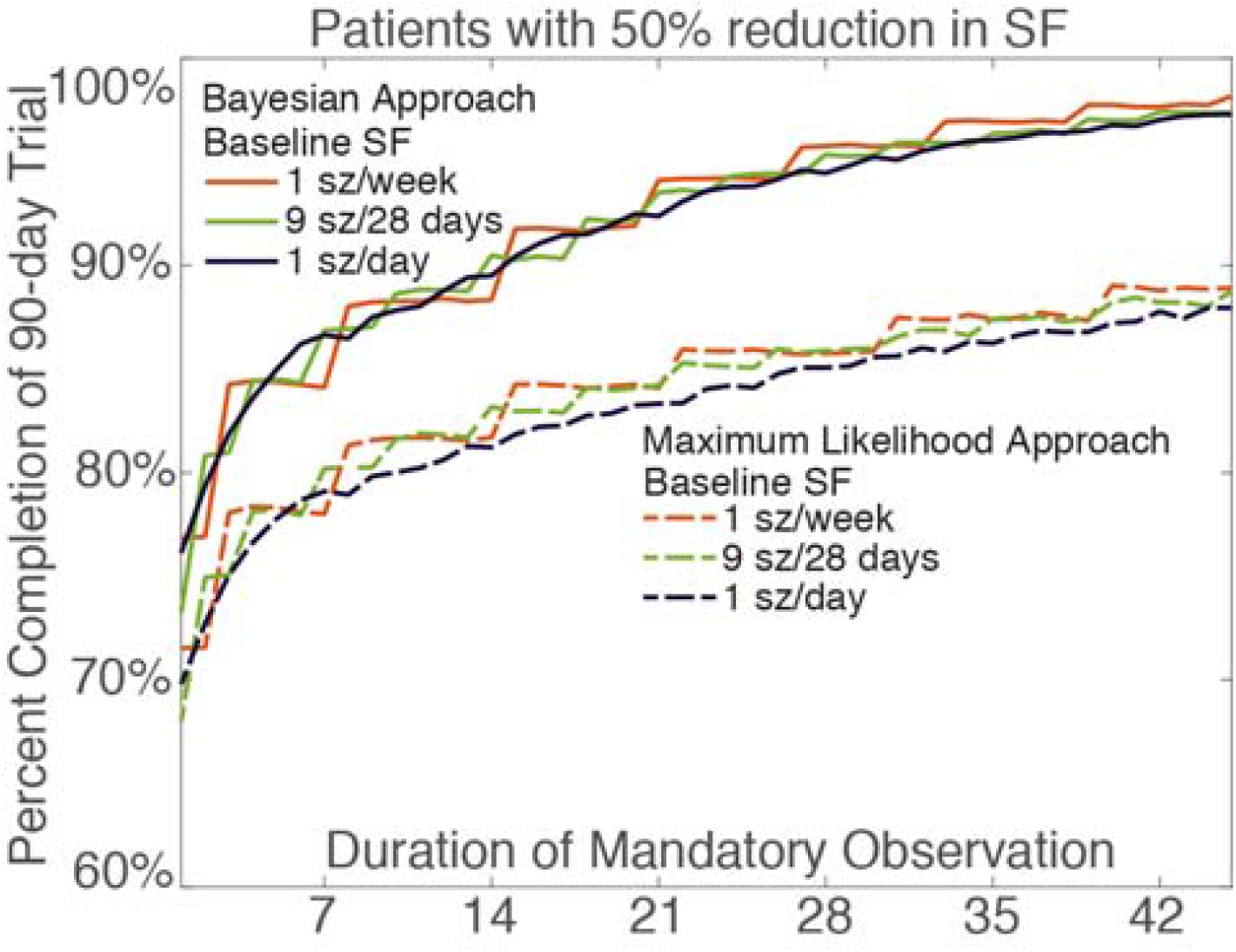
Percent retention in the trial for 100,000 simulated participants who had a 50% reduction in seizure frequency (SF) with treatment based upon the mandatory minimum observation period. The disjointed nature of the 4 seizures per 28 baseline days reflects the transition of the threshold for nonresponse at one day to the next highest integer.

The probabilities that participants had greater than 50% of their baseline seizures during the treatment period if their SF had reduced by 50% using both methods is illustrated in Supplemental Table 2.

### 3.2 Response-adaptive crossover randomization simulations

Compared to canonical trials, there were improvements in statistical power using both of the response-adaptative trial designs except when comparing placebo to the treatment that reduced SF only 25% (Figure 5). The cutoff of the number of enrolled participants to achieve 80% power to detect the actual difference between treatments with a false positive rate of 5% is displayed in Table 3. Variation in these cutoffs was estimated using bootstrapping and the width of 95% bootstrap intervals over 10,000 bootstraps was only 3 participants, therefore these tight intervals are not displayed. In comparing the two response-adaptive trial designs, the BPP method resulted in substantially less time in less efficacious treatment arms with a corresponding increase in the time in the most effective treatment arm (Figure 6).

**Figure 5:**
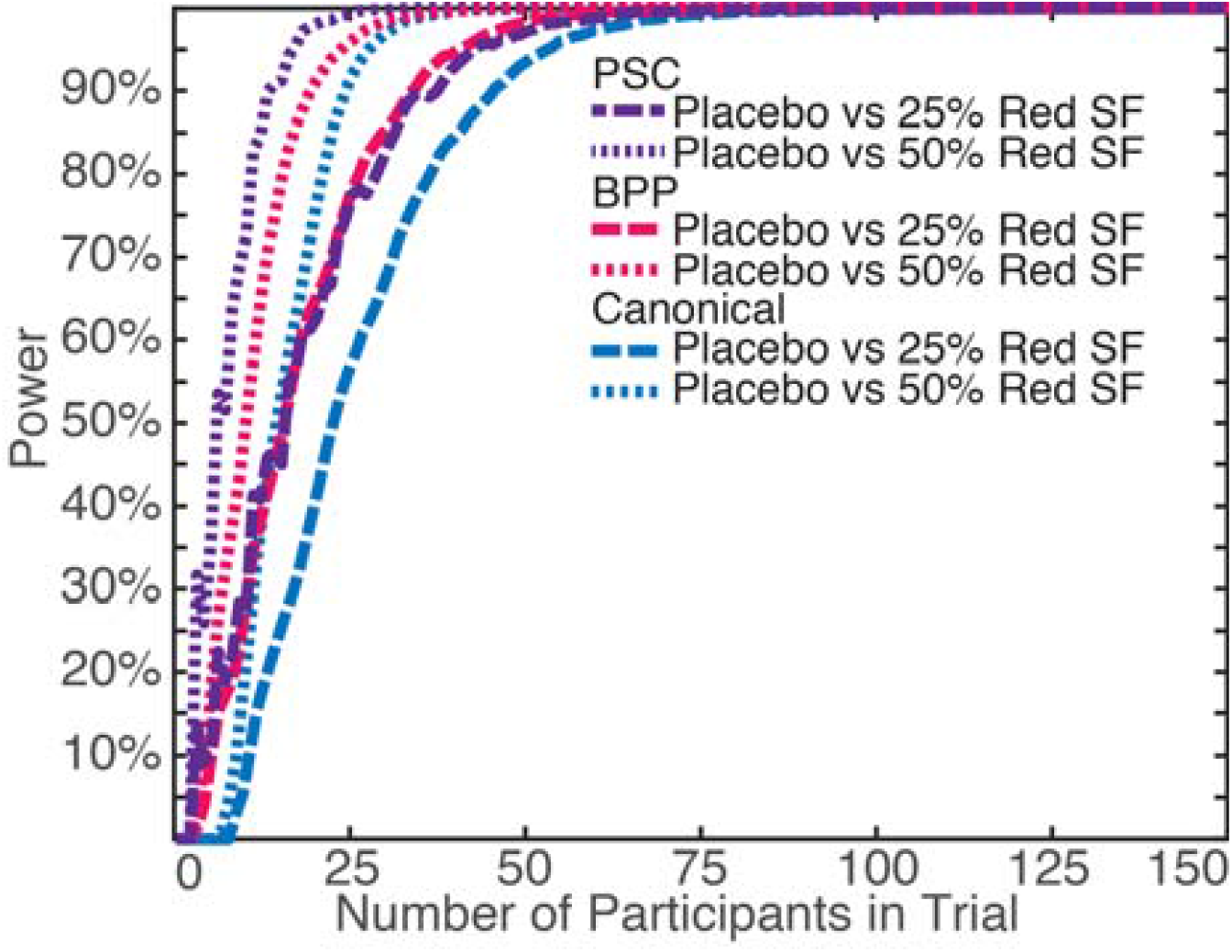
Statistical power based on sequential participant enrollment to detect differences between three treatments that result in 0% (placebo), 25% and 50% reduction in seizure frequency (Red SF) compared to a prerandomization baseline of 9 seizures per 28 days using a canonical trial design, or response-adaptive crossover design using a time to PSC or BPP criterion for response.

**Figure 6:**
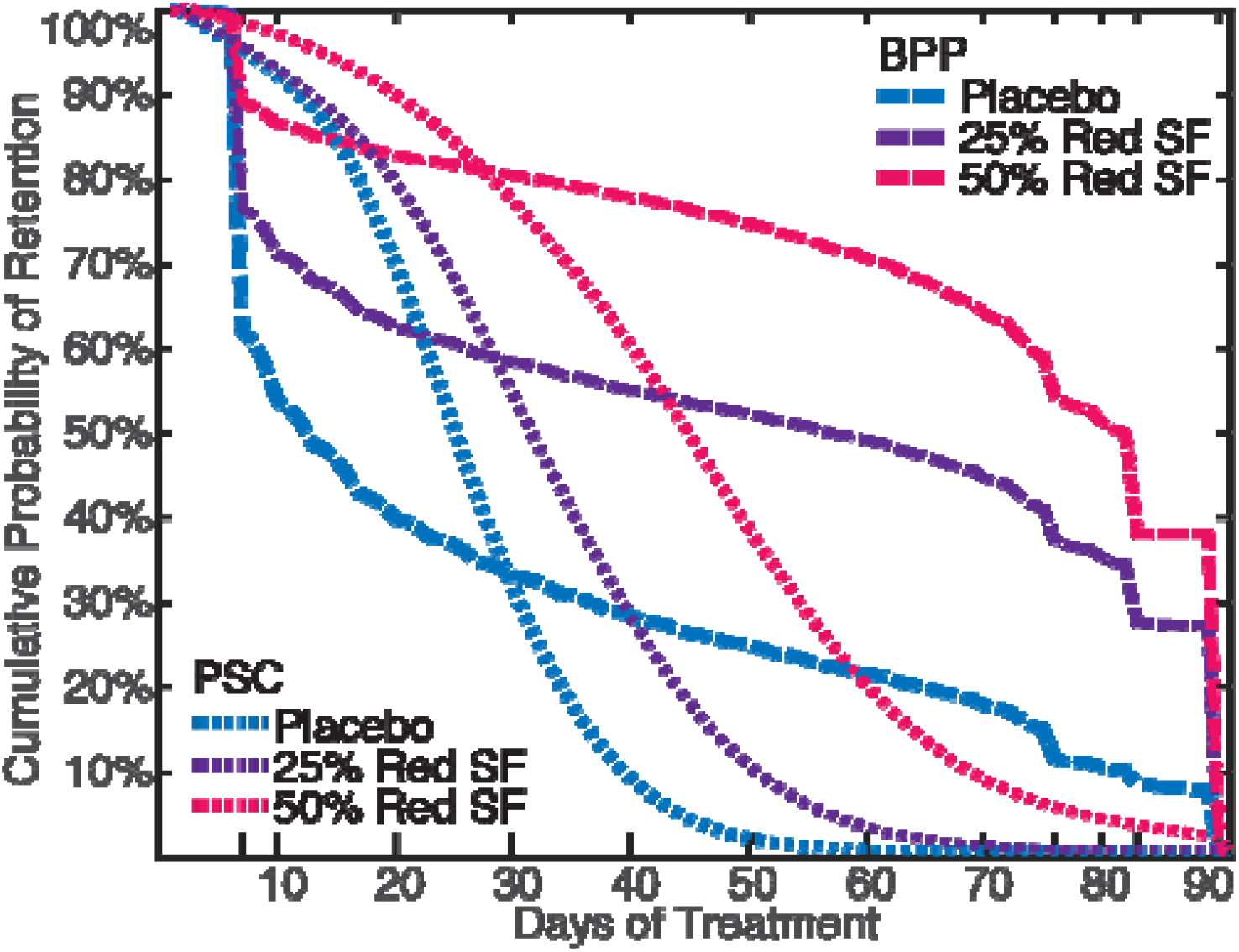
Probability of retention in the treatment arm using a BPP criterion for exit compared to time to PSC for participants with 9 seizures per 28-day baseline. Analogous to Figure 3B. Discontinuity at 76 and 83 days reflect participants initially randomized to other treatments. Abbreviations: see Figure 5.

## 4. Discussion

This manuscript illustrates a novel approach to design of a randomized controlled clinical trial to simultaneously maximize the statistical power of a trial while also maximizing the amount of time participants receive the most efficacious treatment.

Consider an illustrative example where a participant has a PSC of 7 (1 seizure per 4 days). The participant logs 2 seizures during the first treatment week, which is more than expected (1.75 seizures) but not high enough to determine a non-response (p=11% by MLE and 8% by BPP) or worsening of seizures (p=22% by MLE). If this participant has 2 seizures each during the second and third weeks, the total is 6 in 21 days. At that time the probability that the participant’s seizure frequency has halved is 4% for the MLE approach and 2.7% for the Bayesian approach. Using PSC, the participant would wait for 1 more seizure before re-randomization, but a canonical trial could continue this treatment for 69 additional days during which the patient would be expected to have 19 more seizures. To explore the Poisson approach using a variety of baseline and treatment seizure frequencies, please refer to our online calculator: https://wesleykerr.shinyapps.io/PoissonSeizureFrequency/.

The simplified rule of thumb that this analysis suggested is that if the participant had greater than 75% of their baseline seizure frequency during any duration of treatment period greater than 1 week, the participant had less than a 5% chance of halving their baseline SF, as long as 5 to 10 seizures were observed.

Compared to a canonical trial, both the time to PSC and Poisson modeling resulted in modest reductions in the number of participants required to achieve 80% power (Table 3, ^6^). The marginal improvement in power in PSC was difference likely was due to more balanced sampling. Using PSC, participants spent less time on the most efficacious treatment, resulting in increased exploration of the less efficacious treatments (Figures 5 and 6, ^18^). Once a participant was assigned to the treatment that resulted in a 50% reduction in seizure frequency, the PSC criterion resulted in less than 5% of patients completing 90 days, whereas the BPP criterion resulted in 87% 90-day completion. This difference highlights that both the seizure count and the amount of time needed to achieve that seizure count are important to determining non-response to treatment.

If the simultaneous goals of a clinical trial is to demonstrate efficacy of the treatment, maximize the safety of the participants through reduction in SUDEP and other risks from continued seizures, and monitor for treatment-related adverse effects primarily in the most efficacious treatment ^3^, then this approach may serve as a substantial improvement from standard practice. However, there are multiple practical limitations that we did not explore in this initial theoretical framework.

First, this approach supposes that participants log their seizures accurately every day and, if they were determined to not respond, have immediate access to the subsequent treatment assignment. While the accuracy of participant-reported seizure frequency is unpredictably variable ^1^, to switch treatments using a response-adaptive design, one would need to reveal the treatment assignment and perform real-time analysis of results. Double blinding then would necessitate automated processing of results and treatment assignments.

Secondly, a Poisson process approximates the statistical distribution of seizures, but there are many currently unmodeled sources of variation that limit the direct application of this model. Participants are more motivated to enroll in experimental treatments after their seizures have worsened, therefore artificially increasing their measured baseline seizure frequency, with subsequent regression towards their lower, long-term baseline seizure frequency irrespective of treatment ^19^. Further, placebo treatment does reduce seizure frequency, which would result in longer time in the placebo arm and reduced power to detect how treatment changes seizure frequency, controlling for placebo response ^20; 21^. Additionally, within a trial there are titration and pharmacokinetic effects of medications. Seizures that do or do not occur during titration offer valuable information but incorporating them into the model relies on knowledge of the dose-response curve and the central-nervous system effective dose at the time the seizure occurred based on the treatment’s half-life and steady state concentration ^16^.

Outside of clinical trials, long term monitoring through SeizureTracker™ suggests that a 1-day back autoregressive negative binomial model may provide advantages over the simple Poisson model ^7; 9^. The choice of a 1-day back was based on the best fit across 682 adult and 844 pediatric participants ^7^. However, clinical experience suggests that some participants may have more propensity to cluster than others, and therefore may require further modeling to account for individual-level clustering. Longer-term seizure count modeling suggests that there may be weekly, monthly (especially in some menstruating female participants), and potentially annual trends that influence seizure frequency ^8^. Further, the responsive neurostimulator (RNS) data suggests that participants and caregivers can consistently under or over-count seizures in particular participants, which may be only partially controlled for by comparing to the individual-participant’s baseline seizure count ^1^. This long yet incomplete list of unmodeled factors may be why negative binomial distributions better model real-world seizure diary data, because a negative binomial is similar to an overdispersed Poisson ^7^. Due to these considerations, our simple Poisson is a utilitarian framework that may be adapted by modeling non-Poisson sources of variation.

As with most statistics, observing more seizures (data) allowed for better estimation of the baseline and treatment SF and reduced misidentifications. The Bayesian approach is more conservative than the MLE approach is because it incorporates uncertainty in the baseline seizure frequency from observing a finite number of seizures. Individual-level modeling of non-Poisson contributions to variation in seizure count would likely require more pre-randomization and treatment monitoring.

With the understanding of these limitations in using a Poisson process, the general approach for earlier estimation of likelihood allows for reduction of the exposure time to an ineffective treatment and therefore benefits the participant’s safety while maintaining a similar power to the PSC approach, and offering improvements in power relative to the canonical approach. This statistical approach allows for a possible individual-level results in weeks instead of months. This analysis also suggests that the previous threshold of excluding subjects with a doubling SF may expose participants to unnecessary risk from requiring participants to experience additional seizures that did not impact the final conclusion of the trial.

Even though a short observation period can determine efficacy, trials also simultaneously evaluate for the presence of adverse effects to medications. While adverse effects tend to occur soon after dose initiation ^22^, shorter observation may reduce trials’ power for detection of longer-term side effects (e.g. weight gain) ^6^. The response-adaptive cross-over design allows for observation for adverse effects in treatment arms that are potentially effective, but limits observation in treatment arms that are less effective. In our opinion, if a participant’s seizures were not responding a treatment, then prolongation of ineffective therapy to monitor for additional adverse treatment effects may be unethical. However, if a minimum of 8% of participants assigned to placebo continue to appear to respond to “treatment,” these participants may provide appropriate comparisons to participants on active treatment arms (Table 2). While the power to observe statistical differences in adverse events could be reduced, this reflects the population-level cost-benefit considerations that underlie the design of response-adaptive clinical trials ^4; 5^.

While the efficacy of treatment could be determined quickly, the parameters of the model that can be modified based on the cost-benefit ratio of the specific trial designers. Our practical simulations showed that the false positive rate for determining non-response was high within the first week and reduced asymptotically (Figure 3). When we required observation for at least 1 week, more than 80% of simulated participants with a 50% reduction in seizures completed a 90-day trial, irrespective of the baseline seizure rate. This retention fraction also can be increased by allowing for participants with less response to treatment to remain in that treatment arm (e.g. 25% SF reduction) or reducing the threshold for switching of a less than a 5% likelihood of response (e.g. to α<1%). In general, we caution against making treatment decisions based on observing a single seizure in participants who have infrequent seizures prior to three or more times the baseline inter-seizure interval ^23; 24^.

Consideration of both the number of seizures and the time in which these seizures occurred, relative to baseline, could result in modestly more efficient and markedly safer clinical trials for seizures. Compared to PSC, our approach shortened the duration of less effective treatments while lengthening the duration of more effective treatments, with only a small reduction in overall trial power.

Regarding placebo or treatments that do not change seizure frequency, the median patient with a PSC of 9 was on this ineffective treatment for 16 days using our method, 28 days using PSC, and 90 days in a canonical trial. These 12 to 74 additional days on ineffective treatment may harm the participant. Furthermore, the re-randomization may improve the trial’s ability to characterize effect treatments.

## Data Availability

There are no primary data associated with this work.

## 5. Acknowledgements

This work was supported by NIH R25 NS065723 and the Eisenhower Health Department of Internal Medicine.

## 6. Declaration of Interest Statement

The authors have no competing interests to declare. We confirm that we have read the Journal’s position on issues involved in ethical publication and affirm that this report is consistent with those guidelines.

---

**Supplemental Table 1.** Total participants required to achieve 80% power with each trail design based on 10,000 simulated trials. Abbreviations: false positive rate (α), power (1−β), Prerandomization Seizure Count (PSC); Bayesian Posterior Probability (BPP).

| Total participants for<br>1-β>80% for $\alpha$<5% | BPP | PSC | Canonical |
| --- | --- | --- | --- |
| Placebo vs 25% SF reduction | 28 | 30 | 37 |
| Placebo vs 50% SF reduction | 17 | 12 | 22 |

## Notes

### Competing Interest Statement

The authors have declared no competing interest.

### Author Declarations

There are no data for an IRB/oversight body to approve.

