## Supplemental Methods for "Individual-level response adaptive crossover trial design for epilepsy: structure and simulation"

Supplemental Text

Wesley T. Kerr, Xingruo Zhang, & John M. Stern

The following will demonstrate the statistical basis by which we determine the probability that a patient has either failed or responded to an anti-seizure medication (ASM) based on real-time reporting of their seizures during a clinical trial. This is organized in the following sections:

1. Stating the problem based on statistical assumptions and distributions
2. Illustrating the framework of the solution using conventional hypothesis testing
3. Incorporating uncertainty of estimation into the solution using a Bayesian approach
4. Empiric estimation of the duration of treatment exposure

We first address the question of how to determine the likelihood that the patient has failed a medication. Subsequently, we address the probability of falsely identifying a patient as a non-responder if the threshold of experiencing more than half the baseline seizures is used.

### 1 Statistical Problem Statement and Assumptions

First, we model the statistical process that generates seizure counts, $N$, over a certain period of time, $t$, as Poisson processes in which the following assumptions are made:

1. The probability of a seizure during a given period of time, $t$, is proportional to the length of that interval.
$$P[N(t) > 0] \propto t$$
2. Two seizures cannot occur simultaneously.
3. The probability of a seizure does not change unless medication changes are made.
4. The probability that a seizure occurs during one time interval is independent of if a seizure occurred during a non-overlapping time interval.

While most of those assumptions likely are true for seizures, there are some key situations where they are violated. Our approximation of seizure counts as a Poisson process is only as good as those assumptions hold true. For more detailed discussion of the benefits and risks of these assumptions, please refer to the main text.

Based on those properties of a Poisson process, we then determine that the probability that patient has $n$ seizures during time interval $t$ is given by:

$$P[N(t) = n|\lambda] = \frac{(\lambda t)^n}{n!} e^{-\lambda t}.$$

In a clinical trial with a baseline period, we can find the maximum likelihood estimate of the baseline $\lambda$, denoted as $\lambda_0$, for each patient with the following formula:

$$\lambda_0 = \frac{n_{\text{baseline}}}{t_{\text{baseline}}}.$$

In a clinical trial where we are assessing if a patient responds to an ASM, we define a response to an ASM as a 50% reduction in seizure frequency. In our notation, that would be equivalent to halving $\lambda_0$. Therefore, in the treatment phase, we would like to evaluate, given our reported seizure count, $n_t$, the probability that $\lambda_t$ is less than or equal to half of $\lambda_0$.

Alternatively, this approach of looking for non-response can be modified seamlessly to looking if the patient hasn't worsened or have experienced an at least doubling of seizure frequency. To evaluate if the patient hasn't worsened, the threshold of $\lambda_t$ is less than or equal to half of $\lambda_0$ should be changed to testing if $\lambda_t$ is less than or equal to $\lambda_0$ itself. To test for doubling, we first calculate the probability that $\lambda_t$ is less than or equal to two times $\lambda_0$ then subtract that probability from 1 to estimate the probability that $\lambda_t$ is greater than two times $\lambda_0$.

#### 2 Framework of the Solution

Therefore, for a given seizure count during the treatment period, $n_t$, we would like to determine the probability that seizure count, or higher, would occur if the rate of seizures had at least halved. This corresponds to a null hypothesis that the seizure rate is $\lambda_t \leq \lambda_0/2$ and a one-tailed alternative hypothesis that $\lambda_t > \lambda_0/2$. We write the probabilistic expression as:

$$P\left[N(t) \geq n_t \,\middle|\, \lambda_t \leq \frac{\lambda_0}{2}\right].$$

If we take the worst case scenario that the seizure rate had just halved, we recognize this as the right sided cumulative distribution function of the Poisson distribution, which is the upper incomplete gamma function, $\Gamma\left(n_t, \frac{\lambda_0 t}{2}\right)$. This logic is illustrated in the following expression:

$$P\left[N(t) \geq n_t \,\middle|\, \lambda_t \leq \frac{\lambda_0}{2}\right] \leq \Gamma\left(n_t, \frac{\lambda_0 t}{2}\right).$$

Therefore, based on the reported seizure frequency during the baseline period, we can create a table of situations where this probability is less than 5%, suggesting that the null hypothesis that seizure frequency has been halved is violated with a false positive rate, $\alpha$, of 5%. This table is illustrated in the main text as Table 1.

As a point of discussion, if the actual new seizure rate, $\lambda_t$, was less than half of $\lambda_0/2$ then the actual seizure rate would be lower, but the number of seizures that needed to be observed prior to determining that the patient had not responded to the new treatment would not change. In this situation, the probability of enough seizures occurred to reject the null hypothesis would be lower than 5% but, because we did not know that the actual $\lambda_t$, we would have to consider the worst cast scenario that $\lambda_t = \lambda_0/2$.

Secondly, this same framework can be used to address how often patients with seizure frequencies less than half the baseline seizure frequency report seizure counts higher than half the baseline rate during the treatment period. This expression is as follows:

$$P\left[N(t) > \frac{n_0}{2} \middle| \lambda_t \leq \frac{\lambda_0}{2}\right] = \Gamma\left(\lceil \frac{n_0}{2} \rceil, \frac{\lambda_0}{2}\right).$$

Where $\lceil$ and $\rceil$ are the ceiling operators and the left side of the equation models $N(t) > n_0/2$ instead of $N(t) \geq n_t$.

### 3 Incorporating statistical uncertainty

One complication behind the simple solution described above is that in the above solution, we utilized the maximum likelihood estimate of $\lambda_0$ and $\lambda_t$. Next, we consider the possibility that the actual estimated baseline seizure rate, $\hat{\lambda}$, is different from this maximum likelihood estimate, and then repeat the process illustrated above. We incorporate this uncertainty by utilizing a weighted contribution of the possible options for $\lambda_0$ where the weights are based upon the probability of that $\lambda_0$ given the observed number of seizures in the baseline period, $n_0$.

To determine these probability-based weights, we use Bayes formula as below:

$$P\left(\lambda_0 = \hat{\lambda} \,|N(t) = n_0\right) = \frac{P\left(N(t) = n_0 \middle| \hat{\lambda} = \lambda_0\right) P\left(\lambda_0 = \hat{\lambda}\right)}{\sum_{\forall \lambda} P\left(N(t) = n_0 \,|\lambda\right)}. \tag{1}$$

We estimate the prior distribution of $\lambda$, $P\left(\lambda_0 = \hat{\lambda}\right)$, using a uniform distribution across all possible values for $\lambda_0$ ranging from one seizure every 28 days to 24 seizures per day for 28 days in a row in steps of one seizure every 28 days. The shape of this probability-based weighting for example values of $n_0$ is illustrated in the main text as Figure 1.

We also can use this structure to determine the likelihood of each seizure rate, $\lambda_t$, during the treatment phase based on the reported seizures during the treatment phase, $n_t$. In this way, we can assess the likelihood that $2\lambda_t \leq \lambda_0$ if the patient reports $n_t$ seizures using the following formula:

$$P\left(2\lambda_t \le \lambda_0 \,\middle|\, \begin{array}{c} N(t)=n_t \\ N(0)=n_0 \end{array}\right) = \sum_{\forall\lambda_0} P(\lambda_0 \,|N(0)=n_0\,)\, P\left(2\lambda_t \le \lambda_0 \,\middle|\, \begin{array}{c} N(t)=n_t \\ \lambda_0 \end{array}\right)$$
$$= \sum_{\forall\lambda_0} P(\lambda_0 \,|N(0)=n_0\,) \sum_{\forall 2\lambda_t \le \lambda_0} P(\lambda_t \,|N(t)=n_t\,)\,.$$

We highlight that instead of calculating the probability of $N(t) \ge n_t)$ as we did with conventional hypothesis testing, we are instead calculating $P(2\lambda_t \le \lambda_0 | N(t) = n_t, N(0) = n_0)$. Because the Bayesian methodology, it is not necessary to consider $P(2\lambda_t \le \lambda_0)|N(t) \ge n_t, N(0) = n)$. This is because the integral necessary to calculate a cumulative distribution function for a p-value are calculated in reference to $2\lambda_t \le \lambda_0$ as compared to in reference to $N(t) \ge E(n_0/2)$.

For the first term of that expression, we use the Bayesian solution in equation (1) above. Similarly, for the second term of the expression, we also use a Bayesian solution as follows:

$$P\left(2\lambda_t \le \lambda_0 \,\middle|\, \begin{array}{c} N(t)=n_t \\ N(0)=n_0 \end{array}\right) = \sum_{\forall\lambda_0} P(\lambda_0 \,|N(0)=n_0\,) \sum_{\forall 2\lambda_t \le \lambda_0} \frac{P(N(t)=n_t \,|t\lambda_t\,)\, P(\lambda_t = \lambda)}{\sum_{\forall\lambda} P(N(t)=n_t \,|t\lambda\,)}\,.$$

We include $t$ in this expression because we are considering the number of seizures, $n_t$, that occur in a certain number of days during the treatment period, which may be different from 28 days.

For the prior distribution of $\lambda_t$, we again use the uniform distribution across the same range as prior. If we had assumed that $\lambda_t$ was unchanged by treatment, this would bias our analysis towards concluding that the seizure rate during the treatment period, $\lambda_t$, was unchanged from the baseline period, $\lambda_0$.

We can determine the minimum $n_t$ for which this probability is less than 0.05 to determine the number of seizures needed to be observed in a specified treatment period to determine that the patient has not responded to the treatment. A table of these minimum seizure counts, $n_t$, is displayed in the main text as Table 2.

Secondly, to use this similar Bayesian framework for the question of falsely identifying patients as non-responders when they report more than half as many seizures as they did during the baseline period. This corresponds to the following statistical expression:

$$P\left[N(t) > \frac{n_0}{2} \,\middle|\, \lambda_t \le \frac{\lambda_0}{2}\right] = \sum_{\forall\lambda_0} P(\lambda_0 \,|N(0)=n_0\,)\, \Gamma\left(\lceil \frac{n_0}{2} \rceil, \frac{\lambda_0}{2}\right).$$

### 4 Estimating the Duration of Observation to Determine Non-Response

To determine how long patients' would be exposed to treatment using our approach, we simulate patients' seizures with Poisson random walks. We presume that patients will update seizure count daily, therefore, we create the Poisson random walk by decomposing the up-to-90-day into a sum of 90 independent Poisson random variables. We remind the reader that due to the first and fourth properties of a Poisson process (listed above), a 90-day Poisson process is statistically identical to the sum of 90 independent 1-day Poisson proccesses with $\lambda_{1\text{ day}} = \frac{\lambda_{90\text{ day}}}{90}$. In these random walks, after each day we ask if the number of seizures observed is higher than the minimum number of seizures to determine non-response for that patient's baseline seizure frequency, $\lambda_0$.

For each possible $\lambda_0$ from 1 to 100 seizures in the baseline 28-days and for a range of fractional change in seizure frequency from an 80% reduction to a 100% increase (doubling) in steps of 10%, we simulated 10,000 independent patients using both the maximum likelihood estimate and the Bayesian posterior probability estimate. From these simulated patients we built an empiric probability distribution of the number of observation days to determine non-response. The number of simulated patients, 10,000, was chosen based on the canonical rule of thumb for permutation tests that to determine cutoffs for $\alpha = 5\%$ ordinal statistics, roughly that number of simulations is needed for stability of cutoff. For illustration, we chose $\lambda_0$ corresponding to the minimum number of seizures in the 28-day baseline, 4, the rough median of previous trials for antiseizure medication, 10, and a high but reasonable rate of one seizure per day.
